# A Cannabidiol/Terpene Formulation Increases Restorative Sleep in Insomniacs: A Double-Blind, Placebo-controlled, Randomized, Crossover Study

**DOI:** 10.1101/2023.06.03.23290932

**Authors:** Michael Wang, Marcus Faust, Scott Abbott, Vikrant Patel, Eric Chang, John I. Clark, Nephi Stella, Paul J. Muchowski

**Author notes:** Corresponding Author: Paul J. Muchowski, Ph.D. President, Defined Research Institute; 1250 Missouri Street Unit #312 San Francisco, CA 94107. **Author Declarations** All listed authors have seen and approved this manuscript. Funding for this study was provided by Defined Research, Inc. M.W., M.F., S.A., V.P., E.C. and P.J.M. hold equity in Defined Research Inc., a for-profit company. J.C. and N.S. are unpaid consultants who hold stock option grants for equity in Defined Research Inc. **Clinical Trial Registration** ClinicalTrials.gov NCT05233761.

## Abstract

**Study Objectives:** Cannabidiol (CBD) is increasingly used as a health supplement, though few human studies have demonstrated benefits. The primary objective of this study was to evaluate the effects of an oral CBD-terpene formulation on sleep physiology in insomniacs.

**Methods:** In this double-blind, placebo-controlled, randomized clinical trial, 125 insomniacs received an oral administration of CBD (300 mg) and terpenes (1 mg each of linalool, myrcene, phytol, limonene, α-terpinene, α-terpineol, α-pinene, and β-caryophyllene) for ≥ four days/week over four weeks using a crossover design. The study medication was devoid of Δ^9^-Tetrahydrocannabinol (Δ^9^-THC). The primary outcome measure was the percentage of time participants spent in the combination of slow wave sleep (SWS) and rapid eye movement (REM) sleep stages, as measured by a wrist-worn sleep-tracking device.

**Results:** This CBD-terpene regimen significantly increased the mean nightly percentage of time participants spent in SWS + REM sleep compared to the placebo [mean (SEM), 1.28% (0.60%), 95% C.I. 0.09 to 2.46, *P* = 0.03]. More robust increases were observed in participants with low baseline SWS + REM sleep, as well as in day-sleepers. For select participants, the increase in SWS + REM sleep averaged as much as 48 min/night over a four-week treatment period. This treatment had no effect on total sleep time (TST), resting heart rate or heart rate variability, and no adverse events were reported.

**Conclusions:** Select CBD-terpene ratios may increase SWS + REM sleep, and have the potential to provide a safe and efficacious alternative to over-the-counter (OTC) sleep aids and commonly prescribed sleep medications.

**BRIEF SUMMARY:** *Current Knowledge/Study Rationale:* Physicians are increasingly asked by their patients regarding the merits of using CBD for insomnia and other ailments, but lack any rigorous clinical research to support recommending its use. The current study represents the first double-blind, placebo-controlled and randomized crossover clinical trial to investigate how an oral formulation of cannabidiol (CBD) and terpenes influences sleep physiology in insomniacs.

*Study Impact:* In contrast to many OTC sleep aids and commonly prescribed sleep medicines, the CBD-terpene formulation increased SWS and REM sleep, which are critical for the immune system, tissue regeneration, cognition and memory. These results, if confirmed in larger clinical trials, suggests that CBD might offer a promising alternative to other prescription sleep medications and OTC sleep aids.

## INTRODUCTION

Insomnia is the most common sleep disorder and an established risk factor for anxiety, depression and other diseases. ^1^ Approximately ∼30-40% of the adult population in the United States. alone (∼63-84 million) report symptoms of insomnia, and the prevalence of insomnia increases with age. ^1^ Insomnia is defined clinically as the perception or complaint of inadequate or poor-quality sleep due to a number of factors, such as difficulty initiating or maintaining sleep, waking up too early in the morning, or having nonrestorative sleep. ^1^ It causes significant distress and/or impairment in daytime functioning. ^1^ A clinical diagnosis of insomnia is typically obtained by patient-reported complaints about their sleep. ^2^ Objective testing is not usually recommended unless another disorder is suspected, yet researchers often uses objective testing in sleep studies. ^2^

CBD is a bioactive ingredient produced by *Cannabis* and hemp, and was approved by the Food and Drug Administration (FDA) in 2018 for the treatment of seizures associated with two rare and severe forms of epilepsy, Lennox-Gastaut syndrome (LGS) and Dravet syndrome (DS). ^3^ Preclinical and early human studies suggest that CBD may modulate sleep physiology. A study in rats showed that CBD increases SWS sleep in a dose-dependent manner. ^4^ In a placebo-controlled clinical trial, participants with insomnia (n = 15) that received 160 mg of CBD reported a subjective increase in their sleeping time relative to those in the placebo control arm in a sleep survey, but this study did not track objective measures of sleep physiology. ^5^ In an open label study in adults with clinically diagnosed anxiety (n = 72), CBD (25-75 mg/day) improved sleep quality in ∼66% of the patients as determined by a subjective sleep quality questionnaire. ^6^ Most recently, a randomized controlled pilot trial (n = 15) of CBD (150 mg/night) over a period of two weeks in participants with moderate to severe insomnia showed no effect on subective and objective measures of sleep, but an improvement on participant well-being. ^7^ Significantly, somnolence was the most frequent adverse event reported in previous clinical studies in LGS and DS. ^8^

Terpenes are a class of small molecules produced by most plants, including *Cannabis* and hemp. While we know that select terpenes are sedating in mice, ^9–13^ their effects on sleep physiology in humans have not been studied.

The objective of this interventional study was to evaluate the effects of oral administration of a CBD-terpene formulation on sleep physiology, as measured by a wrist-worn sleep-tracking device over a treatment period of four weeks in 125 participants with insomnia.

## METHODS

### Aims, objectives and hypotheses

The goal of this trial was determine the effects of a oral CBD-terpene formulation on sleep physiology in insomniacs. The primary endpoint of the study was to determine if the CBD-terpenes formulation increases SWS + REM (% TST) sleep over a treatment period of four weeks, as quantified with an objective wrist-worn sleep-tracking device. The null hypothesis was that there was no difference in SWS + REM (% TST) sleep in participants that received CBD-terpenes and the placebo control; the alternative hypothesis was that there was a significant difference among the two treatment groups.

### Study design and participant population

This study involved a double-blind, placebo-controlled, randomized, crossover design in which participants cycled through two independent treatment arms (Group 1 Treatment A: Placebo; Treatment B: CBD-terpenes; Group 2 Treatment A; CBD-terpenes; Treatment B: Placebo), each for four weeks (Fig. 1). The study involved a two-week run-in/baseline period during which participants were monitored for protocol compliance to ensure they were wearing the sleep-tracking device and collecting sleep data correctly. The run-in/baseline period was followed by two independent four-week treatment periods (A/B and B/A), which were each followed by a one-week wash-out period (Fig. 1). A one-week wash-out period was chosen to rule out any carry-over effects, as the elimination half-life of CBD is two-five days after chronic oral administration. ^14^

**FIGURE 1.**
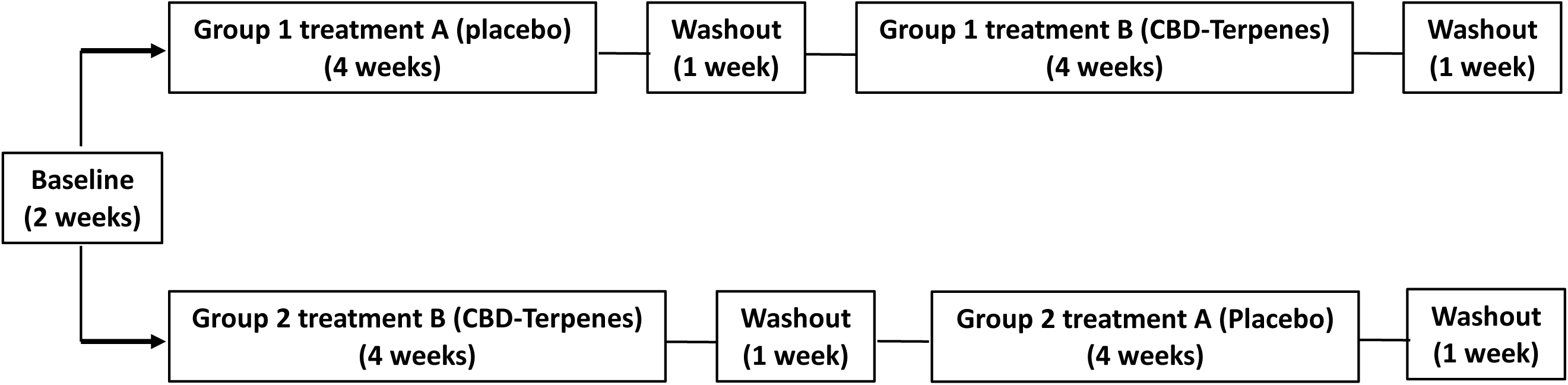
Crossover study design.

This study was approved by an Institutional Review Board (IRB) (Allendale IRB, Lyme, CT). This study was performed in a decentralized manner with participants recruited from across the United States. Recruitment, enrollment, informed consent, and distribution of study materials to participants was managed by a contract research organization (83 Bar, Austin, TX). Recruitment was initiated on 18 February, 2022. 13,385 potential study leads were recruited from across the United States via social media advertising (Fig. 2). After a prospective study participant clicked on a social media ad they were directed to a dedicated landing page that included an embedded survey. The survey was designed to pre-screen prospective candidates. The presence of chronic insomnia in all participants was determined in this online survey as a self-reported difficulty initiating (latency to persistent sleep >30 min) and/or maintaining sleep (>30 min awake during the middle of the night, or waking >30 min before desired waking time on three or more nights per week) for at least three months. In addition, all participants scored as having severe insomnia after taking a clinically validated insomnia severity index ^15^ that was also part of the survey. The study identified 422 qualified leads from survey respondents, who were further screened via phone interviews conducted by registered nurses to ensure that potential study participants met all of the inclusion and exlusion criteria, resulting in the enrollment of 125 participants (101 females and 24 males) in the study (Fig. 2). Each participant provided written informed consent prior to participation. The average insomnia severity index score of participants in our study was 26 ± 2 (severe clinical insomnia) out of a scale from 0-28. ^15^

**FIGURE 2.**
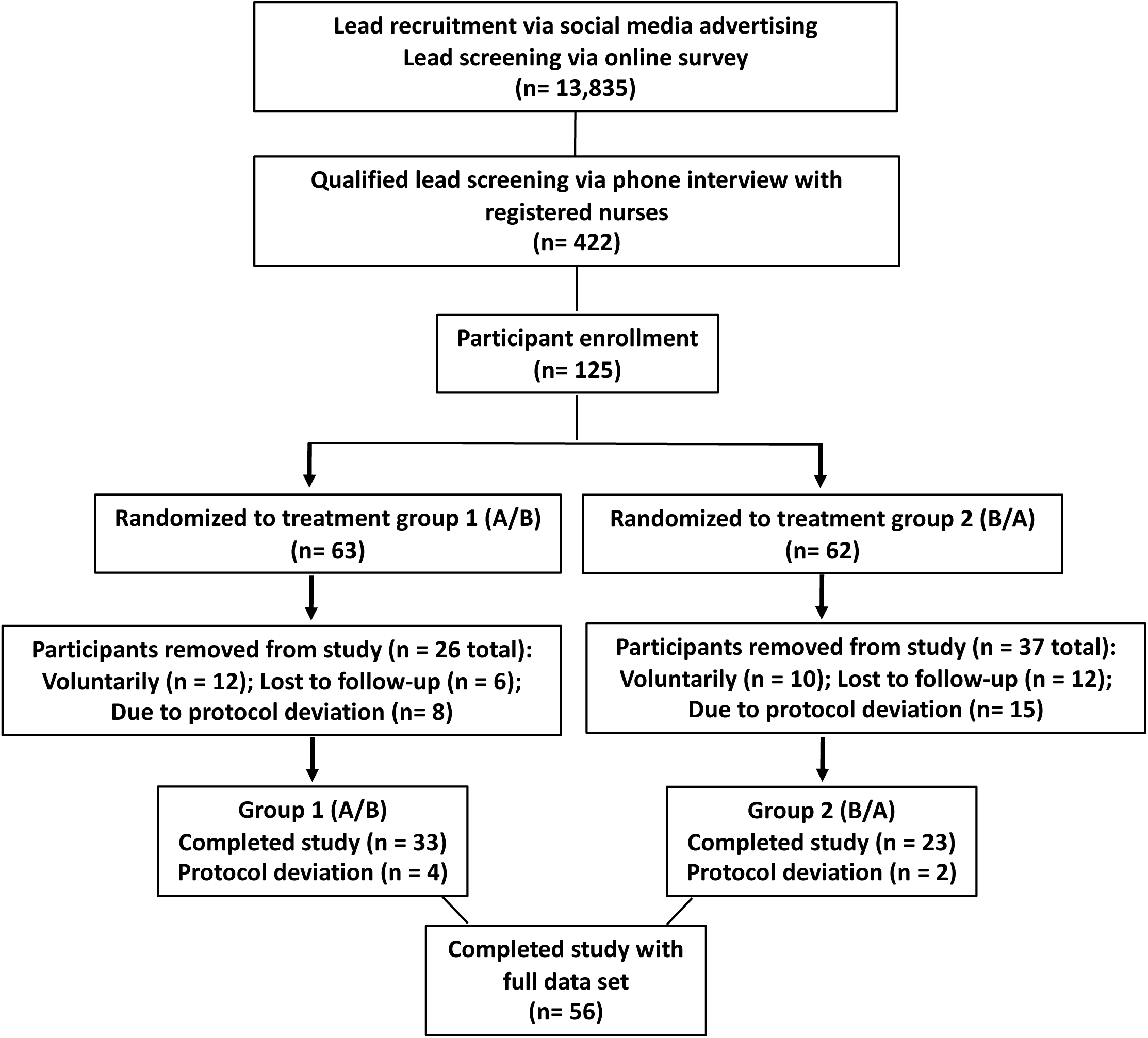
Flow diagram of study participation.

### Allocation and blinding

125 participants with chronic insomnia were randomized in a 1:1 allocation ratio to one of two treatment sequences (n = 63 A/B sequence group and n = 62 B/A sequence group) using a computer-generated randomization code (https://www.randomizer.org) that was implemented by an independent researcher (Fig. 2). Allocation was concealed and medications were numbered to mask the randomization code. Study participants and research personnel were blinded to randomization sequence and group allocation throughout the study. Every participant cycled through both treatment arms. Participants were instructed to take the treatment on a minimum of four nights/week. Throughout the study on a daily basis, participants responded to a text-based survey to determine if they had taken the study medication.

### Study medication

The study medication was composed of capsules made from vegetable cellulose that contained >99.9% purity hemp-derived CBD (300 mg) and one mg each (>98% purity, food grade) of the terpenes linalool, myrcene, phytol, limonene, α-terpinene, α-terpineol, α-pinene, and β-caryophyllene dissolved in organic coconut oil. The CBD and terpene doses were established from a series of in-house dose-ranging studies (data not shown). The CBD and terpenes were purified in good manufacturing practice (GMP)-certified facilities. Participants took capsules with a glass of water one hour before going to sleep. Of note, Δ^9^-THC was undetectable in the study medication, as measured by an independent ISO-10725-accredited analytical testing laboratory (Supplementary Data Figure 1). Furthermore, capsules were free from potential contaminants (pesticides, residual solvents or heavy metals) (data not shown).

Placebo-control capsules were identical to those that contained the treatment medication, but contained only organic coconut oil.

### Efficacy Assessments

The study was conducted in a decentralized manner in which all participants took treatments and slept in their own homes. Objective sleep data was gathered throughout the study from a non-invasive sleep-tracking wrist-worn device (https://www.whoop.com) that electronically collected and transmitted sleep data from study participants in the comfort of their own beds. The wristband collects hundreds of data points per second from a three-axis accelerometer, three-axis gyroscope, and heart rate sensor. The wristband can accurately measure TST (min) and the time spent in sleep stages [defined as Light sleep, SWS (Deep) sleep, REM sleep, and Awake]. The device also collects data using photoplethysmography (PPG), a technology that quantifies blood flow by measuring superficial changes in blood volume. Heart rate, heart rate variability and respiratory rate are derived from PPG data, and these metrics are processed by Whoop’s sleep detection and staging algorithms. Two recent validation studies, published independently from Whoop, indicate that data on sleep stages collected from the Whoop device correlate well with polysomnography (PSG), the gold-standard of sleep tracking used in clinical studies conducted in sleep clinics. ^16,17^ Specifically, in one study the Whoop device had low bias (13.8 minutes) and precision (17.8 minutes) errors for measuring sleep duration, and measured REM sleep and SWS accurately (intraclass coefficient, 0.74 ± 0.28 and 0.85 ± 0.15, respectively). ^16^ In addition, the accuracy of Whoop-based measurements of heart rate, respiratory rate, and heart rate variability were excellent compared with the gold-standard PSG. ^16^ In a second independent study that compared the Whoop device to PSG, the sensitivity (i.e., the percentage of PSG-determined sleep epochs correctly identified by the Whoop device) to light sleep, SWS, and REM sleep was 62%, 68%, and 70%, respectively. ^17^

Participants were blinded to the sleep data collected by the Whoop device, and responded to a daily text-based questionnaire to report throughout the duration of the study whether or not they had taken the study medication on the previous night. Thus, objective sleep data were analyzed only for nights when participants took the study medication.

### Questionnaires

A modified version of a clinically validated questionnaire/survey entitled the patient’s global impression (PGI) ^18^ was used to assess subjective perceptions of sleep by the study participants after each four-week treatment period. The PGI is a four-item, subjective, participant self-report that assesses treatment benefit to sleep quality (Item 1), sleep induction (Item 2), sleep duration (Item 3), and appropriateness of study medication strength (Item 4). ^18^ Each item in the PGI is presented as a survey to patients that consists of a three-point categorical scale, with a score of 1 representing a treatment benefit/advantage on items 1 to 3 (“too strong” on item 4), a score of 2 representing no effect/change on items 1 to 3 (“just right” on item 4), and a score of 3 representing worsening/disadvantage on items 1 to 3 (“too weak” on item 4). We modified the PGI by removing Item 4 (appropriateness of study medication strength) from the survey. Our modified PGI also included two additional items to assess a potential treatment benefit to sleep depth (item 4), and treatment effect on the presence of vivid dreams (Item 5). Moreover, each item in our modified PGI was presented as a survey that consisted of a ten-point categorical scale, with scores between 6-10 representing a treatment benefit/advantage, a score of 5 representing no effect of treatment, and a score of 1-4 representing a treatment worsening/disadvantage. This data was collected from study participants in the form of a five-min survey that was completed on their mobile phone at the end of each four-week treatment period.

### Safety Assessments

A medical monitor (JGB Biopharma, Belmont, CA) was responsible for assessing adverse events (AEs) for causality and severity, and for final review and confirmation of accuracy of event information and assessments. An AE was defined as any untoward medical occurrence in a study participant during the trial period. A serious adverse event (SAE) was defined as an event that resulted in death, was life-threatening, required patient hospitalization, resulted in a persistent or significant disability/incapacity, and/or resulted in a medically important event or reaction. The following guidelines were used to describe the severity of Aes using the CTCAE (Common Terminology Criteria for Adverse Events) grading system: Grade 1 Mild – Events that required minimal or no treatment and did not interfere with the participant’s daily activities; Grade 2 Moderate – Events that resulted in a low level of inconvenience or concern with the therapeutic measures. Moderate events may cause some interference with functioning; Grade 3 Severe – Events that interrupted a participant’s usual daily activity and may have required systemic drug therapy or other treatment. Severe events are usually potentially life-threatening or incapacitating. The term “severe” does not necessarily equate to “serious”; Grade 4 Life Threatening – Events in which urgent medical intervention was indicated; Grade 5 Death – If related to the event. The medical monitor was responsible for assessing the relationship to study treatment using clinical judgement and the following considerations: Not related: There was not a reasonable possibility that the administration of the study intervention caused the AE, there was no temporal relationship between the study intervention and event onset, or an alternate etiology had been established; Related: The AE was known to occur with the study treatment, there was a reasonable possibility that the study intervention caused the AE, or there was a temporal relationship between the study intervention and event. Reasonable possibility means that there is evidence to suggest a causal relationship between the study intervention and the AE. Participants in our trial were instructed to respond to a daily text-based survey to determine if they had taken the study treatment on the previous evening, and if they had experienced any AEs. In the event that the participant reported an AE, the medical monitor was instructed to follow up directly with the participant to assess the AE for causality and severity.

### Power analysis

Based on the results from two smaller clinical trials we conducted with the CBD-terpenes formulation, we calculated that a total of 100 participants would provide at least 90% power to detect a 15% difference in the percentage of time participants spend in SWS + REM sleep, as quantified using the Whoop sleep-tracking device after the four-week treatment phase. Based on our previous clinical studies, we anticipated a dropout rate of 33%.

### Statistics

The primary endpoint of the study was to determine if a CBD-terpenes formulation increases SWS + REM (% TST) sleep, as quantified with an objective wrist-worn sleep-tracking device. The null hypothesis was that there was no difference in SWS + REM (% TST) sleep in participants that received CBD-terpenes and the placebo control; the alternative hypothesis was that there was a significant difference among the two treatment groups. The mean nightly difference in SWS + REM sleep (% TST) in participants that received CBD-terpenes and the placebo control at the end of the two four-week treatment phases was analyzed using a two-sided test with an alpha level at 0.05 to evaluate superiority. The *P*-value and 95% confidence interval for the estimate of treatment difference in percentages was estimated and constructed using the above-mentioned method.

Objective sleep data was also computed for treatment effects, period effects and carryover effects by the method reported by Hills and Armitage for two-period crosssover clinical trials. ^19^ A pre-test was used to assess potential carryover effects, whereby the sum of the values in the two periods was calculated for each subject and compared across the two sequence groups. If results were statistically significant (*P* < 0.05), then there was evidence of relevant carryover effects and further between groups difference tests were not undertaken. Statistical analysis was performed with GraphPad Prism 10.0.2 (San Diego, CA).

## RESULTS

### Participants

A total of 13,835 potential participants were recruited via social media advertising and screened for isomnia via an online survey (Fig. 2). This resulted in 422 potential participants that qualified for further lead screening via a phone interview with registered nurses, and 125 participants that fulfilled the inclusion criteria and were randomly allocated to the active or placebo group.

During the first two-week baseline period, 63 participants were excluded from the study, either voluntarily (n = 22), lost to follow-up (n = 18) or due to deviation from the study protocol as they did not collect sleep data using the Whoop device (n = 23) (Fig. 2). This high and unanticipated attrition rate resulted in a modified intent-to-treat/per-protocol approach and resulted in 62 participants completing the study. Furthermore, data from six participants [four participants from Group 1 (A/B) and two participants from Group 2 (B/A)] were excluded from the final data analysis due to protocol deviations (e.g. due to a lack of reporting whether or not they took the study treatment, and/or insufficient sleep data collection). Thus, the final data analysis included complete data sets from 56 study participants [33 participants from Group 1 (A/B) and 23 participants from Group 2 (B/A)] (Fig. 2).

### A CBD-terpene formulation increases SWS + REM sleep (%TST) in insomniacs

Study participants self-administered (p.o.) capsules that contained CBD (300 mg) and eight individual terpenes (1 mg each), or a placebo control capsule, on a minimum of four nights/week over a four-week treatment period (Fig. 1). After a one-week washout period, participants crossed over and self-administered the second treatment (i.e., capsules with CBD-terpenes or the placebo control), for a second four-week treatment period (Fig. 1).

Objective sleep physiology data was collected from a wrist-worn sleep-tracking device. ^16,17^ The CBD-terpenes treatment significantly increased the mean nightly time participants spent in SWS + REM sleep (% TST) and SWS sleep (% TST), and decreased the mean nightly light sleep time (%TST), when compared to the placebo control (Fig. 3A, 3B, 3G, and Table 1, respectively). The CBD-terpenes treatment also increased the absolute mean nightly time participants spent in SWS + REM (min) and SWS sleep (min) (Fig. 3D and 3E and Table 1). However, the CBD-terpenes treatment did not change the mean relative (% TST) or absolute (min) time participants spent in REM sleep (Fig. 3C and 3F, and Table 1), nor did it change the mean absolute time (min) participants spent in light sleep (Fig. 3H and Table 1). The treatment had no signficant effect on TST (min) (Fig. 3I and Table 1). No significant effect of sex was observed for any outcome measure (data not shown).

**FIGURE 3.**
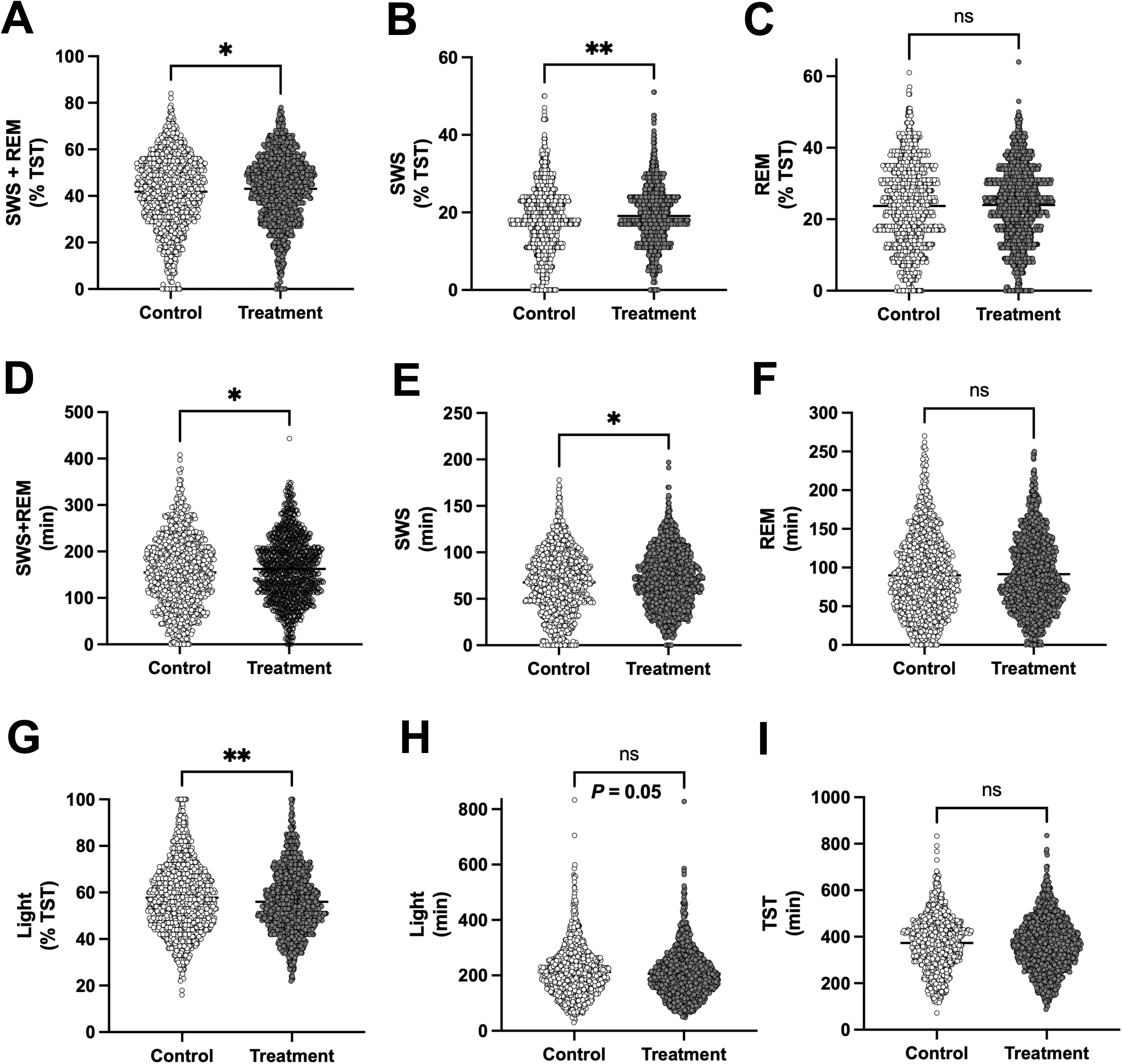
Treatment of study participants with CBD-terpenes significantly increased SWS + REM (%TST) sleep in insomniacs. Treatment with CBD-terpenes increased SWS + REM (% TST) (**3A**), SWS (% TST) (**3B**), SWS + REM (min) (**3D**) and SWS (min) sleep (**3E**), and decreased Light sleep (% TST) (**3G**). This treatment did not change REM (% TST) (**3C**), REM (min) (**3F**), and Light sleep (min) (**3H**), nor did it change TST (min) (**3I**). Shown is the mean nightly value (mean ± SEM) for each outcome measure over a treatment period of 28 days [\**P* < 0.05; ** *P* < 0.01 (t test); n = 56 study participants; note that participants were required to take the treatments ≥ four times/week, and averaged 22 days of treatment for each treatment arm over the course of the 28-day treatment period].

**Table 1:** Mean difference in SWS+REM (% TST), SWS (% TST), REM (% TST), Light (% TST), SWS+REM (min), SWS (min), REM (min), Light sleep (min) and TST.

|  |  |  |  |  |  |  |  |  |  |
| --- | --- | --- | --- | --- | --- | --- | --- | --- | --- |
|  | All Study Participants<br>(n = 56) |  |  | Low Baseline SWS+REM<br>(%TST) (n = 21) |  |  | Day-sleepers (n = 7) |  |  |
| Outcome Measure | MD<br>(SEM) | 95%<br>CI | P | MD<br>(SEM) | 95% CI | P | MD<br>(SEM) | 95% CI | P |
| SWS+REM (% TST) | 1.3 (0.6) | 0.1 to 2.5 | * | 3.7 (0.9) | 2.0 to 5.5 | **** | 8.3 (2.0) | 4.4 to 12.1 | **** |
| SWS (% TST) | 1.0 (0.3) | 0.4 to 1.6 | ** | 1.3 (0.5) | 0.4 to 2.3 | ** | 3.7 (0.9) | 1.9 to 5.5 | **** |
| REM (% TST) | 0.3 (0.4) | -0.6 to 1.2 | n.s. | 2.4 (0.7) | 1.0 to 3.7 | *** | 3.4 (1.5) | 0.5 to 6.3 | * |
| Light (% TST) | -1.7 (0.6) | -2.8 to -0.5 | ** | -3.7 (0.9) | -5.5 to -2.0 | **** | -8.1 (2.0) | -11.9 to -4.2 | **** |
| SWS+REM (min) | 7.5 (3.0) | 1.7 to 13.3 | * | 10.2 (4.3) | 1.8 to 18.7 | * | 19.7 (10.0) | 0.0 to 39.3 | * |
| SWS (min) | 3.3 (1.3) | 0.7 to 6.0 | * | 2.7 (2.0) | -1.2 to 6.6 | * | 10.2 (3.7) | 2.9 to 17.5 | ** |
| REM (min) | 1.3 (2.0) | -2.7 to 5.3 | n.s. | 7.2 (3.0) | 1.3 to 13.0 | * | 12.5 (6.7) | -0.6 to 25.7 | n.s. |
| Light (min) | -1.7 (0.6) | -2.8 to -0.5 | * | -16.8 (6.7) | -30.0 to -3.6 | * | -34.9 (12.0) | -55.3 to -11.3 | ** |
| TST | -6.9 (4.7) | -16.0 to 2.4 | n.s. | -19.0 (8.5) | -35.7 to -2.3 | * | -11.3 (13.8) | -38.4 to 15.8 | n.s. |
| Shown is the mean difference between control and treatment arms for the total population of study participants (n = 56), participants with low baseline SWS+REM (%TST) (n = 21), and day-sleepers (n = 7). *$P < 0.05$, **$P < 0.01$; ***$P < 0.001$; ****$P < 0.0001$ (t test). Participants were required to take the treatments ≥ four times/week, and averaged 22 (total participants), 21 [low baseline SWS+REM (%TST)], and 25 (day-sleepers) days of treatment for each treatment arm over the course of the 28-day treatment period. | | | | | | | | | |

Objective sleep data was also computed for treatment effects, period effects and carryover effects by the method reported by Hills and Armitage for two-period crosssover clinical trials. ^19^ This analysis confirmed that the CBD-terpenes treatment significantly increased SWS + REM sleep (% TST) and SWS sleep (% TST), and decreased light sleep (% TST), when compared to the placebo control (Supplementary Table 1). No evidence of relevant carryover or period effects was observed for the CBD-terpenes treatment on these outcome measures (data not shown). Using this statistical analysis, the CBD-terpenes treatment did not significantly change REM sleep (% TST), nor did it change the absolute time participants spent in SWS + REM (min), SWS (min), or REM (min) sleep, although there was a trend towards increasing absolute SWS + REM sleep (min) (Supplementary Table 1). The CBD-terpenes treatment did not significantly change TST (min) (Supplementary Table 1). No significant effect of sex was observed for any outcome measure (data not shown).

Analysis of the entire SWS + REM (% TST) data set from the study, suggested that the CBD-terpenes treatment may have skewed the data distribution towards a decrease in the time participants spent in low levels (0-24%) of SWS + REM sleep (% TST), and an increase participants spent in high levels (25-54%) of SWS + REM sleep (% TST) (Supplementary Fig. 2A). To assess, retrospectively, if there was an association between treatment with CBD-terpenes and SWS + REM (% TST) sleep, this data was analyzed as a contingency table. A Fisher’s exact test confirmed that treatment with CBD-terpenes signficantly shifted the distribution of data away from low levels and towards high levels of SWS + REM sleep (% TST) (*P* = 0.004; Odds ratio 1.428; 95% C.I. 1.124 to 1.810) (Supplementary Fig. 2A). Conversely, analysis of the entire Light sleep (% TST) data set from the study, suggested that the CBD-terpenes treatment may have skewed the data distribution towards an increase in the time participants spent in low levels (0-54%) of Light sleep (% TST), and a decrease participants spent in high levels (55-100%) of Light sleep (Supplementary Fig. 2B). To assess, retrospectively, if there was an association between treatment with CBD-terpenes and Light sleep (% TST), this data was analyzed as a contingency table. A Fisher’s exact test confirmed that treatment with CBD-terpenes signficantly shifted the distribution of data away from high levels and towards low levels of Light sleep (% TST) (*P* = 0.048; Odds ratio 0.8483; 95% C.I. 0.7231 to 0.9982) (Supplementary Fig. 2B).

### Treatment with CBD-terpenes increases SWS + REM sleep (% TST) and other objective sleep outcome measures in the majority of study participants

For numerous FDA-approved and OTC drugs, the reponse rate to drug treatment in clinical trials is highly variable, due in part to genetic variants in drug metabolism. ^20,21^ In the current study we observed that SWS + REM sleep (% TST and min) increased in 55% and 61% of study participants, respetively, treated with CBD and terpenes (Table 2). In contrast, light sleep (% TST and min) decreased in 57% and 61% of study participants, respectively (Table 2). For some objective outcome measures evaluated in this study, including SWS + REM (% TST), SWS (% TST) and SWS + REM (min), the percentage of participants that responded positively to treatment with CBD-terpenes was ∼2-fold higher than those with a negative response, and ∼4-fold higher than those with a neutral response (Table 2). For comparison, clinical studies of CBD treatment in children with LGS and DS (for which CBD is FDA approved), indicate that 32% and 43% of patients, respectively, have a positive response to drug treatment. ^22,23^

**Table 2:** Relative percentage of study participants who showed an increase, decrease or no effect of treatment with CBD-terpenes on objective sleep outcome measures.

|  |  |  |  |  |  |  |  |  |  |
| --- | --- | --- | --- | --- | --- | --- | --- | --- | --- |
|  | All<br>Participants<br>(n = 56) | Low<br>Baseline<br>SWS+REM<br>(%TST)<br>(n = 21) | Day-sleepers<br>(n = 7) | All<br>Participants<br>(n = 56) | Low<br>Baseline<br>SWS+REM<br>(%TST)<br>(n = 21) | Day-sleepers<br>(n = 7) | All<br>Participants<br>(n = 56) | Low<br>Baseline<br>SWS+REM<br>(%TST)<br>(n = 21) | Day-sleepers<br>(n = 7) |
| Outcome Measure | Increase | Increase | Increase | Decrease | Decrease | Decrease | No Effect | No Effect | No Effect |
| SWS+REM (% TST) | 55% | 67% | 86% | 30% | 19% | 0% | 14% | 14% | 14% |
| SWS (% TST) | 54% | 52% | 86% | 32% | 33% | 14% | 14% | 14% | 0% |
| REM (% TST) | 48% | 67% | 86% | 38% | 19% | 0% | 14% | 14% | 14% |
| Light (% TST) | 32% | 24% | 0% | 57% | 67% | 86% | 11% | 9% | 14% |
| SWS+REM (min) | 61% | 62% | 86% | 35% | 33% | 14% | 4% | 5% | 0% |
| SWS (min) | 48% | 43% | 86% | 48% | 57% | 14% | 4% | 0% | 0% |
| REM (min) | 50% | 71% | 86% | 48% | 29% | 14% | 2% | 0% | 0% |
| Light (min) | 39% | 29% | 0% | 61% | 71% | 100% | 0% | 0% | 0% |
| TST | 57% | 52% | 43% | 41% | 43% | 57% | 2% | 5% | 0% |

### A subset of study participants show robust beneficial responses to treatment with CBD-terpenes

This clinical study utilized a crossover design in which each participant served as their own control, and therefore the data from each study participant could be quantified individually [equivalent to single-patient (n-of-1) clinical trials]. ^24^ Analysis of the top five responding participant’s sleep data individually (5/56 or ∼9% of all study participants) showed that the CBD-terpenes treatment robustly increased the relative and absolute mean nightly time these participants spent in SWS + REM sleep (% TST and min, respectively) (Supplementary Figs. 3 and 4, and Table 3). In these participants, the nightly mean increase in SWS + REM sleep (min) was 48 min over a four-week treatment period (Supplementary Fig. 4 and Table 3). These participants also averaged 32 min less light sleep per night over the four-week treatment period (data not shown). Of note, here too, the CBD-terpenes treatment did not affect on TST (min) (data not shown).

**Table 3:** Mean difference in SWS+REM (% TST) and SWS+REM (min) for top responding individual study participants (n = 5)

|  |  |  |  |  |  |  |
| --- | --- | --- | --- | --- | --- | --- |
|  | SWS+REM (% TST) | SWS+REM (% TST) | SWS+REM (%TST) | SWS+REM (min) | SWS+REM (min) | SWS+REM (min) |
|  | MD (SEM) | *P* | 95% CI | MD (SEM) | *P* | 95% CI |
| DR24 | 9.1 (3.8) | * | 1.2 to 16.9 | 48.3 (20.1) | * | (7.0 to 89.5) |
| DR45 | 7.6 (2.9) | * | 1.6 to 13.5 | 48.5 (18.6) | * | (11.2 to 85.9) |
| DR51 | 12.7 (3.8) | ** | 4.9 to 20.5 | 51.3 (14.2) | *** | (22.6 to 80.0) |
| DR111 | 5.0 (2.3) | * | 0.3 to 9.6 | 40.5 (14.3) | ** | (11.5 to 69.5) |
| DR128 | 17.3 (3.4) | **** | 10.6 to 24.4 | 48.4 (19.8) | * | (8.3 to 88.4) |
| Shown is the mean difference between control and treatment arms for top responding individual study participants.<br>* *P* < 0.05; ** *P* < 0.01; *** *P* < 0.001; **** *P* < 0.0001 (t test). Participants were required to take the treatments ≥ four times/week, and averaged 19 days of treatment for each treatment arm over the course of the 28-day treatment period. |  |  |  |  |  |  |

### Study participants with low baseline SWS + REM (% TST) sleep (≤ 40%/night) show robust beneficial responses to treatment with CBD-terpenes

The mean nightly SWS + REM sleep (% TST) in hundreds of thousands of users of the Whoop sleep-tracking device is ∼42%. ^25^ We next analyzed the effects of the CBD-terpenes treatment on objective sleep outcome measures in a subset of study participants with low baseline SWS + REM sleep (% TST) (arbitrarly chosen as ≤ 40%/night). In these participants (n = 21/56 or 38% of study participants), treatment with CBD-terpenes increased the mean nightly SWS + REM sleep (% TST) and other objective sleep readouts by a greater magnitude and higher level of statistical significance than in the total study population (Supplementary Fig. 5, Table 1 and Supplementary Table 2). Surprisingly, these beneficial responses were observed despite these participants having potentially lower TST (Supplementary Fig. 5I). Morever, in this subset of study participants with low baseline SWS + REM sleep (% TST), we observed that the relative percentage of participants who responded positively to treatment with CBD-terpenes increased relative to the response rate observed in the total study population (67% in participants with low baseline SWS + REM (% TST) sleep vs. 55% in total study population), while the relative percentage of participants who responded negatively decreased (19% in participants with low baseline SWS + REM (% TST) sleep vs. 30% in total study population) (Table 2).

### Study participants who sleep during the day show the most robust beneficial responses to treatment with CBD-terpenes

One of the exclusion criteria in this study was if any prospective participant was a night shift worker during the 12 months prior to the study, and during the study. Nonetheless, inspection of the raw sleep data indiated that a subset of participants who were enrolled in our study slept only during the daytime. In this subset of study participants (7/56 or ∼13% of study participants), treatment with CBD-terpenes increased mean nightly SWS + REM sleep (% TST) and other objective sleep readouts in a highly significant manner, and by a much greater magnitude than observed in the total study population (Supplementary Fig. 6, Table 1 and Supplementary Table 3). Morever, in these day-sleeping participants, we observed that the relative percentage of participants who responded positively to treatment with CBD-terpenes increased dramatically relative to the response rate in the total study population (86% in day-sleepers vs. 55% in total study population, respectively), while the relative percentage of participants who responded negatively decreased to zero (0% in day-sleepers vs. 30% in total study population) (Table 2).

### Effects of treatment with CBD-terpenes on subjective measures of sleep

A modified version of a clinically validated questionnaire/survey entitled the PGI ^18^ was used to assess insomnia symptoms as perceived by the study participants after each four-week treatment period. This modified PGI is a five-item, subjective, participant self-report that assesses treatment benefit to sleep quality (Item 1), sleep induction (Item 2), sleep duration (Item 3), sleep depth (Item 4), and treatment effect on vivid dreams (Item 5). Treatment with CBD-terpenes modestly improved all of these subjective meaures of sleep in study participants relative to the placebo control, but these improvements did not reach statistical significance (data not shown). Note that participants with low baseline SWS + REM (% TST) and day-sleepers showed a trend to perceive improvements in sleep induction and duration (data not shown), despite their objective data indicating that their TST may have slightly decreased. Remarkably, despite the absence of significant difference in these subjective outcome measures, a higher percentage of study participants perceived a benefit of treatment with CBD-terpenes as opposed to worsening or neutral response when compared to the placebo control treatment (Table 4). Furthermore, the percentage of participants with low baseline SWS+REM (%TST) and day-sleepers who perceived a benefit of treatment with CBD-terpenes to sleep induction, duration and depth was higher than reported by all study participants (data not shown).

**Table 4:**
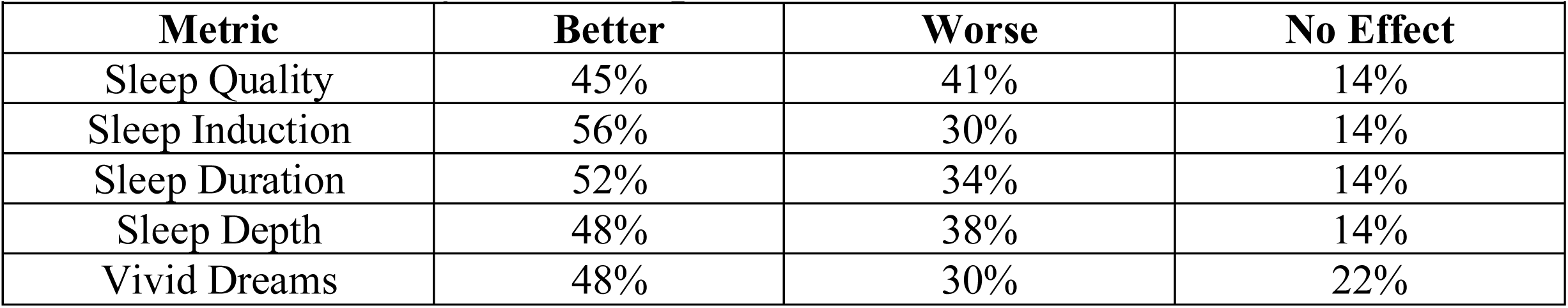
Relative percentage of study participants who showed an improvement, worsening or no effect of treatment with CBD-terpenes on subjective sleep outcome measures (n =56)

### Treatment with CBD-terpenes was well tolerated and was not associated with any adverse events

Study participants responded to a daily text message to describe any potential adverse events they experienced throughout our study, and these text messages were tracked on a daily basis by a medical monitor. No adverse events were reported by any participants throughout this 12-week study; including no significant effect on resting heart rate or heart rate variability (data not shown).

## DISCUSSION

Despite the promising therapeutic potential of CBD and terpenes, sparse human clinical evidence supports their use for the treament of patients with insomnia or other sleep disorders. Here we report that daily (or near-daily) administration of an oral CBD-terpenes formulation significantly increased the mean nightly time that 56 participants with severe insomnia spent in SWS + REM sleep (% TST and min), as measured by an objective wrist-worn sleep tracking device.

The combination of SWS and REM sleep stages is widely referred to as “restorative” sleep, and is thought to strengthen the immune system, increase cell/tissue regeneration and brain metabolite clearance, and replenish energy stores. ^26,27^ SWS and REM sleep are also critical for learning, memory, attention and executive function. ^26,27^ SWS is associated with decreased heart rate, blood pressure, sympathetic nervous activity and cerebral glucose utilization, compared with wakefulness. ^26,27^ Moreover, during SWS, human growth hormone is released while the stress hormone cortisol is inhibited. ^26,27^ The percentage of time spent in SWS and REM sleep may decrease with aging, ^28^ and a decreased perecentage of REM sleep is associated with a greater risk of all-cause, cardiovascular, and other noncancer-related mortality. ^29^ Accordingly, novel and safe therapeutic modalities that selectively increase these sleep stages are urgently needed.

As with many interventional drug studies, we observed variability in the treatment response to CBD-terpenes in our participant population for the objective outcome measures tracked in our study. Given that 55% and 61% of the total study participants had an increase in SWS+REM (% TST and min, respectively), it is not surprising that the magnitude of the treatment benefits across the total study population was relatively modest. Nonetheless, in certain subpopulations in our study (e.g. in participants with low baseline SWS + REM sleep and in day-sleepers), the magnitude of the beneficial responses was much more robust, and achieved higher levels of statistical significance. For example, day-sleepers averaged a nightly increase of 20 min in SWS + REM sleep and a decrease of 35 min in light sleep (Table 1), while the top responding participants in our study averaged a nightly increase of 48 min in SWS + REM sleep (Table 4) over a four-week treatment period. It is worth noting that in previous clinical studies of CBD treatment in children with LGS and DS (for which CBD is FDA approved), only 32% and 43% of patients, respectively, had a positive response to drug treatment. ^22,23^ Numerous studies have shown strong associations between genetic polymorphisms and levels of SWS and/or REM sleep, ^30^ and future studies may elucidate whether such polymorphisms play a role in the response to treatment with CBD-terpenes in insomniacs. There are currently millions of CBD users, and we believe that our results may have important clinical relevance for a reasonable percentage of those individuals, especially those with low SWS + REM sleep, and day-sleepers.

Participants who slept during the day showed the most robust treatment response to CBD-terpenes. We presume that these participants represent night shift workers and may have shift work sleep disorder, a circadian rhythm sleep disorder that causes insomnia and interferes with falling/staying alseep. ^31^ Objective assessment of shift workers by PSG indicates that day sleep is significantly shorter than night sleep, and the sleep loss is primarily taken out of stage two and REM sleep. ^31^ We hypothesize that day-sleepers in our study may efficiently respond to treatment with CBD-terpenes due to lower REM sleep at baseline. Consistent with this scenario, REM sleep (% TST) significantly increased in day-sleepers in the current study (Fig. 7 and Tables 1 and 6), in contrast to our observations in the total study population.

A limitation of the current study is that treatment with CBD-terpenes only modestly improved subjective measures of sleep in study participants relative to the placebo control, and these results fell short of statistical significance. As trends towards statistical significance were observed for some of these subjective outcome measures, we conclude that our study lacked sufficient statistical power to detect such differences, and that future studies should address this shortcoming.

The molecular and systems levels mechanisms that mediate the CBD-terpenes induced increase in SWS and REM sleep will require further studies. One intriguing possibility is that these effects may be mediated by CBD’s modulation of the endocannabinoid system (ECS). For example, CBD has been shown to increase levels of the endocannabinoid anandamide in humans, ^32^ and preclinical studies show that anandamide increases SWS and REM sleep. ^33^

Terpenes are currently classified by the FDA as Generally Regarded As Safe (GRAS) for human consumption and are commonly used as flavoring and aroma additives in food. Preclinical studies indicate that some terpenes are sedating, ^9–13^ but their effects on sleep physiology in humans remains unknown. Numerous studies indicate that terpenes may synergize with cannabinoids to influence physiology by the so-called “entourage effect”, ^34^ but further studies will be required to determine their role in sleep physiology.

Despite tracking by a medical monitor, no adverse events were reported during our trial. These results are consistent with two recent studies documenting a good safety profile for chronic dosing of CBD in humans. ^7,35^ In contrast, adverse events that included dry mouth, dizziness, vertigo, and acute tachycardia were reported by 83% of participants in a previous clinical trial with medical cannabis oil that contained Δ^9^-THC and CBD. ^36^ In addition to these adverse events, Δ^9^-THC decreases REM sleep, ^37^ is intoxicating, and could create fall risks in the elderly. Taken together, our results suggest that select CBD-terpene formulations may be a more effective and safer sleep aid than products which are Δ^9^-THC based. Notably, some commonly prescribed sleep medications and OTC sleep aids induce significant side effects that limit their utility. ^38^ Finally, while many commonly prescribed sleep medications decrease sleep onset latency, they might also decrease SWS and REM sleep, a significant shortfall to these type of medications. ^39^

In summary, our clinical study indicates that combination therapies based on CBD-terpene ratios significantly increases SWS and REM sleep in 56 participants with severe insomnia. This result provides a solid foundation to perform additional placebo-controlled clinical trials with CBD-terpenes in larger cohorts of patients as a potential treatment for insomnnia that increases restorative sleep.

## Data Availability

All data produced in the present work are contained in the manuscript

## ABBREVIATIONS

CBD: cannabidiol
LGS: Lennox-Gastaut syndrome
DS: Dravet syndrome
ECS: endocannabinoid system
PGI: patient’s global impression
REM: rapid eye movement sleep
SWS: slow wave sleep
TST: total sleep time

## ACKNOWLEDGEMENTS

The authors thank Brian Keyes, Dr. Anne Young, Dr. Elise Grenier and Dr. Karen Muchowski for constructive feedback on the mansuscript.

## DATA AVAILABIILTY STATEMENT

The data underlying this article will be shared on reasonable request to the corresponding author.

## AUTHOR CONTRIBUTIONS

MW, MF, SA, VP and EC designed the research studies, conducted experiments and acquired data. JIC and NS designed the research studies and wrote the manuscript. PJM designed the research studies, conducted experiments, acquired data, analyzed data and wrote the manuscript. All authors reviewed and approved the manuscript.

---

**Supplementary Figure 1.**
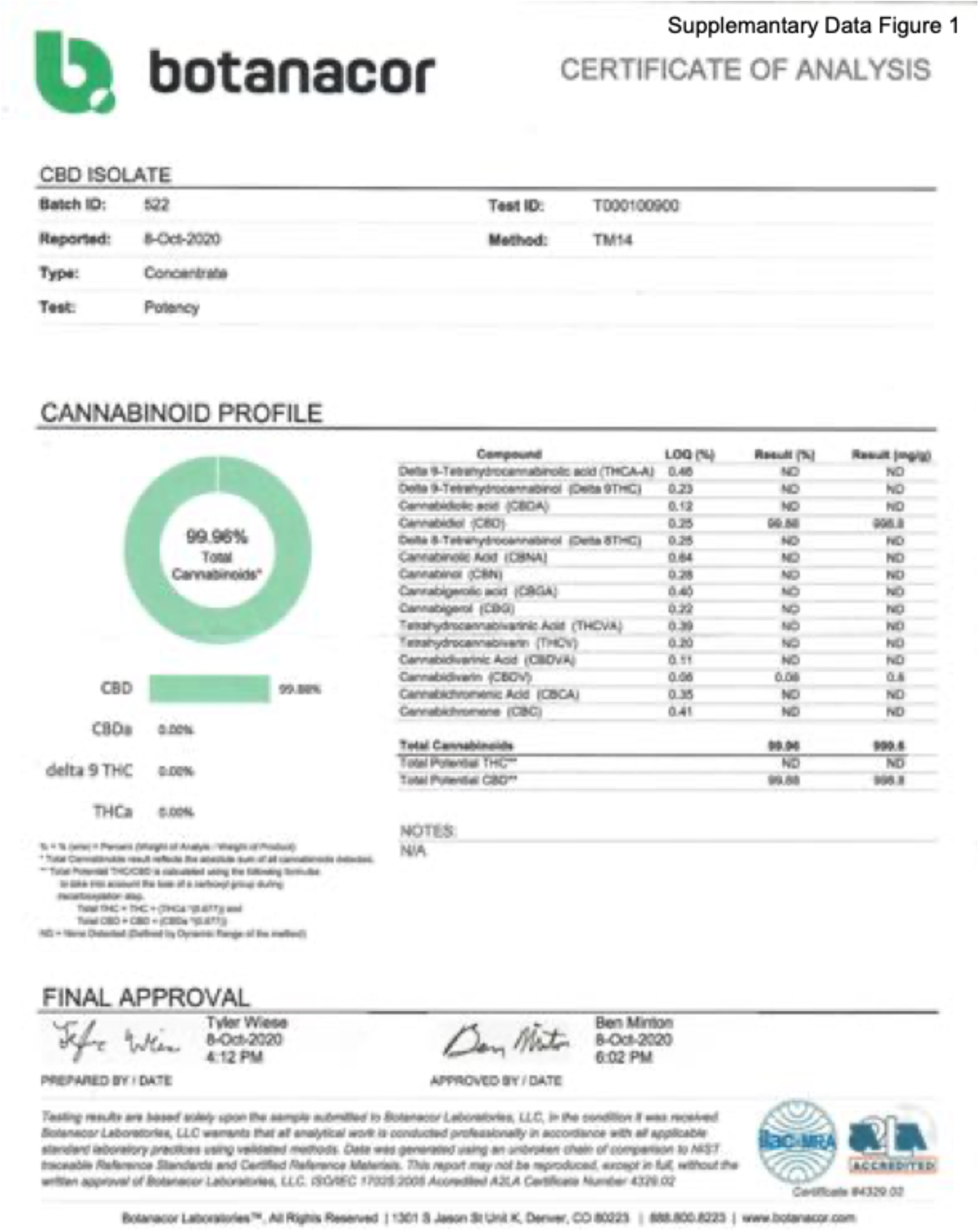

**Supplementary Figure 2.**
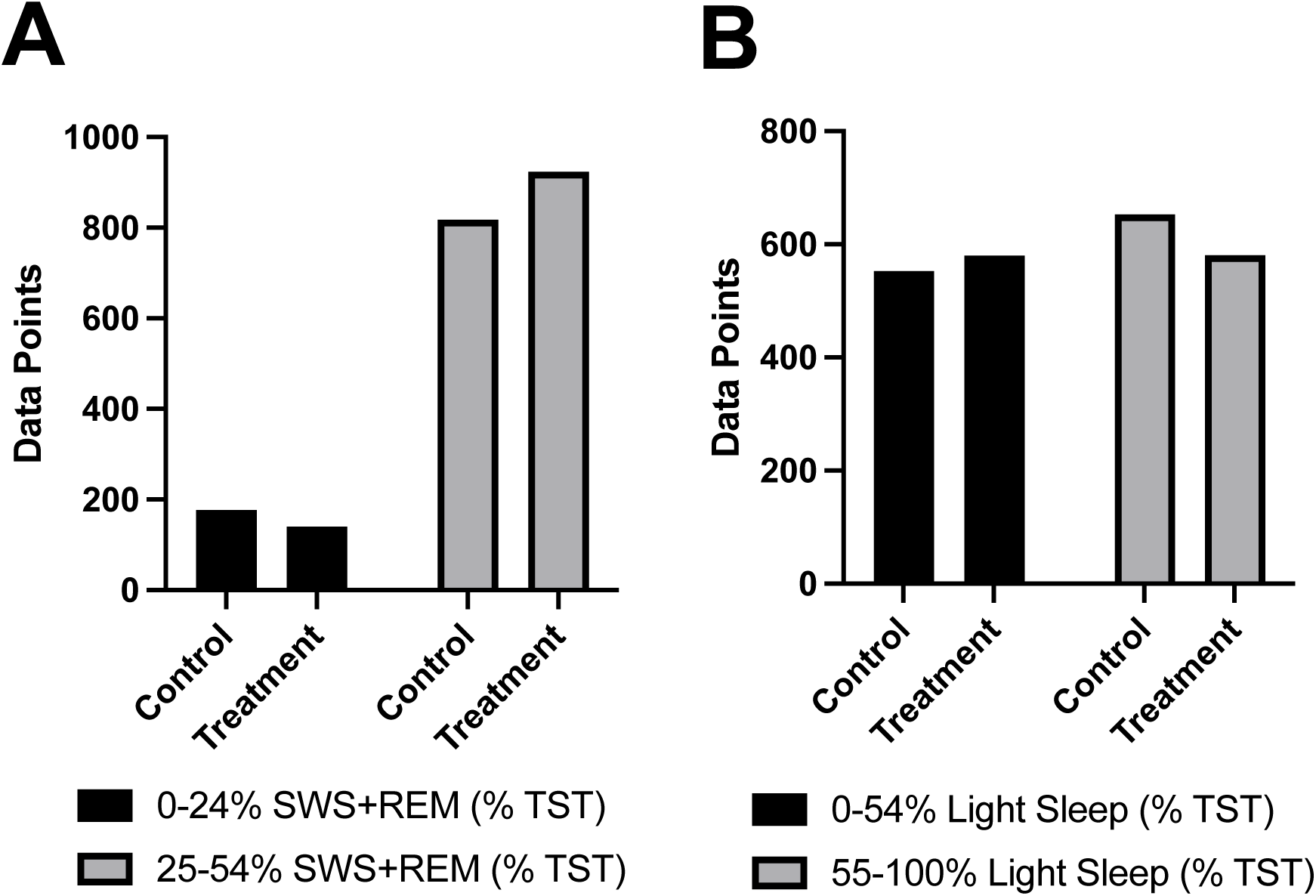

**Supplementary Figure 3.**
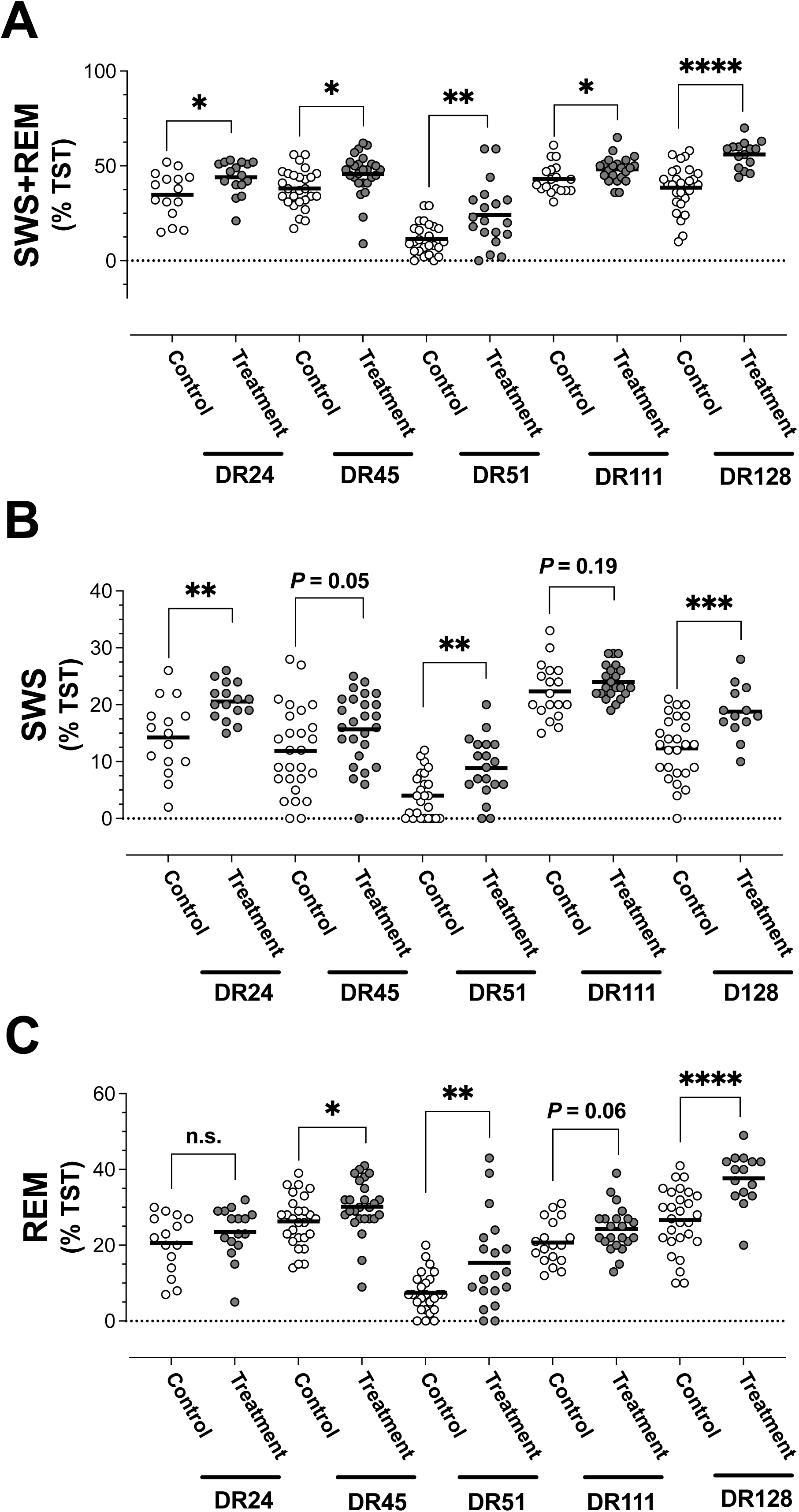

**Supplementary Figure 4.**
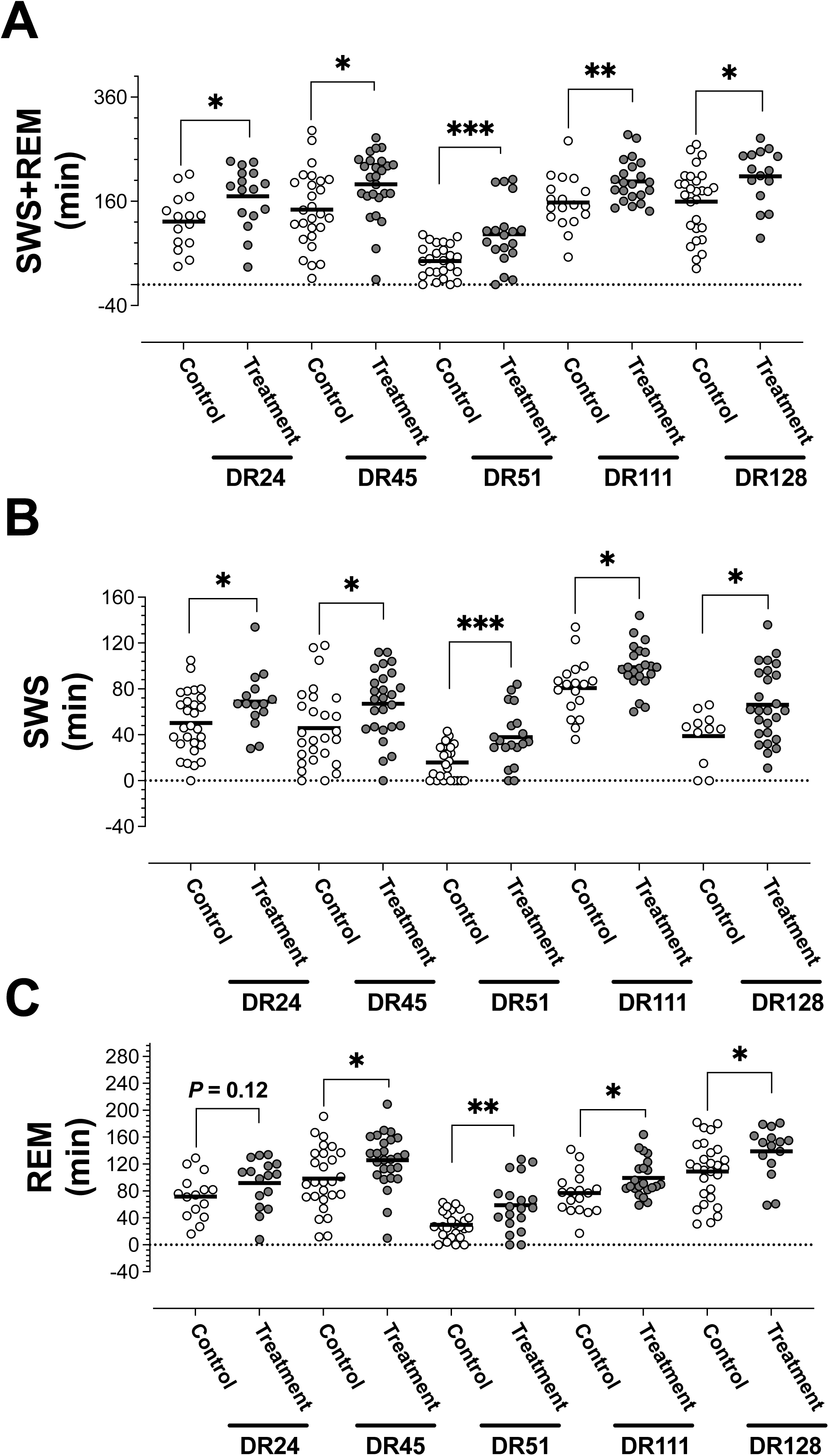

**Supplementary Figure 5.**
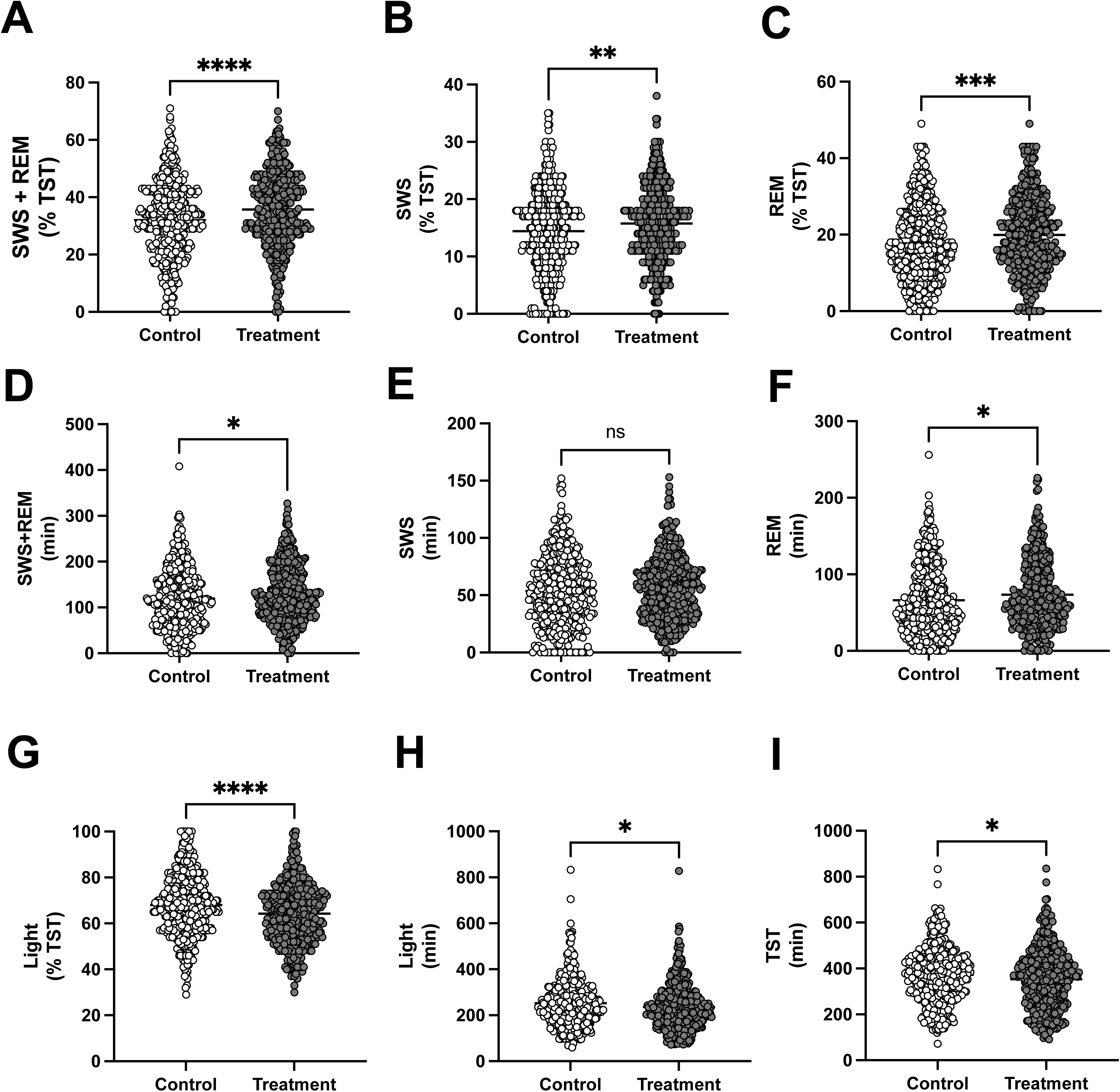

**Supplementary Figure 6.**
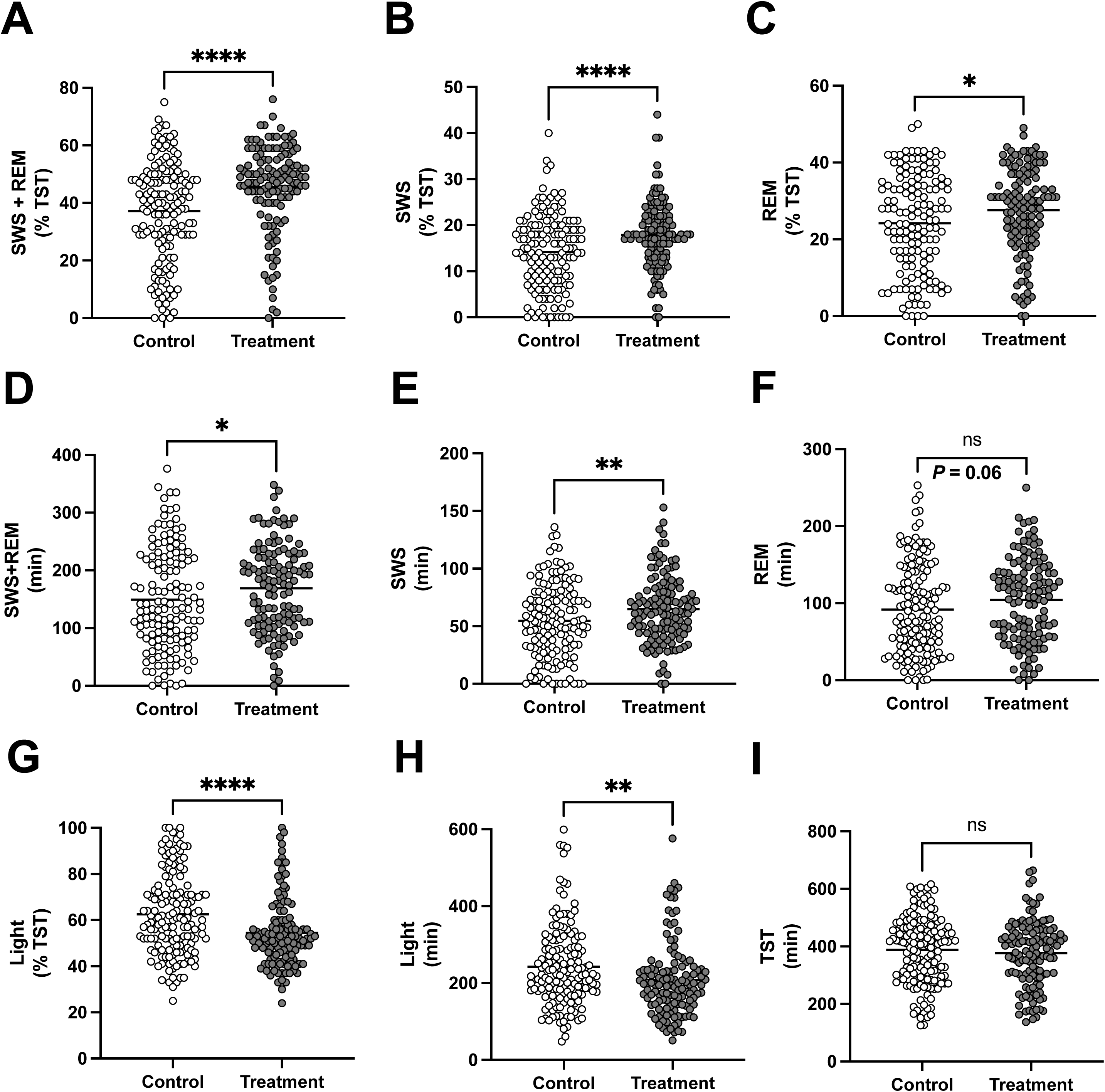

## REFERENCES

1. Black DW, Grant JE, eds. DSM-5 Guidebook: The essential companion to the diagnostic and statistical manual of mental disorders, fifth edition. Washington, DC: American Psychiatric Association Publishing; 2014.

2. Schutte-Rodin S, et al. Clinical guideline for the evaluation and management of chronic insomnia in adults. Clin Sleep Med. 2008; 4(5):487–504.

3. Stella N. THC and CBD: Similarities and differences between siblings. Neuron. 2023; 111(3):302–327.

4. Monti JM. Hypnoticlike effects of cannabidiol in the rat. Psychopharmacology. 1977; 55(3):263–5.

5. Carlini EA, Cunha JM. Hypnotic and antiepileptic effects of cannabidiol. J Clin Pharmacol. 1981; 21(S1):417S–427S.

6. Shannon S, et al. Cannabidiol in anxiety and sleep: A large case series. Perm J. 2019; 23:18–041.

7. Narayan A, et al. Cannabidiol for moderate-severe insomnia: a randomized controlled pilot trial of 150 mg of nightly dosing. J Clin Sleep Med. 2024; Jan 4. doi: 10.5664/jcsm.10998. Online ahead of print.

8. Chesney E, et al. Adverse effects of cannabidiol: a systematic review and meta-analysis of randomized clinical trials. Neuropsychopharmacology. 2020; 45(11):1799–1806.

9. Yang H, et al. α-Pinene, a major constituent of pine tree oils, enhances non-rapid eye movement sleep in mice through GABAA-benzodiazepine receptors. Mol Pharmacol. 2016; 90(5):530–539.

10. Linck VM, et al. Inhaled linalool-induced sedation in mice. Phytomedicine. 2009; 16(4):303–7.

11. Galdino PM, et al. The anxiolytic-like effect of an essential oil derived from Spiranthera odoratissima A. St. Hil. leaves and its major component, β-caryophyllene, in male mice. Prog Neuropsychopharmacol Biol Psychiatry. 2012; 38(2):276–84.

12. Costa JP, et al. Anxiolytic-like effects of phytol: Possible involvement of GABAergic transmission. Brain Res. 2014; 1547:34–42.

13. do Vale TG, et al. Central effects of citral, myrcene and limonene, constituents of essential oil chemotypes from Lippia alba (Mill.) N.E. Brown. Phytomedicine. 2002; 9(8):709–14.

14. Millar SA, et al. A systematic review on the pharmacokinetics of cannabidiol in humans. Front Pharmacol. 2018; 9:1365.

15. Bastien CH, et al. Validation of the Insomnia Severity Index as an outcome measure for insomnia research. Sleep Med. 2001; 2(4):297–307.

16. Berryhill S, et al. Effect of wearables on sleep in healthy individuals: a randomized crossover trial and validation study. Clin Sleep Med. 2020; 16(5):775–783.

17. Miller DJ, et al. A validation study of the WHOOP strap against polysomnography to assess sleep. J Sports Sci. 2020; 38(22):2631–2636.

18. Krystal AD, et al. Long-term efficacy and safety of zolpidem extended-release 12.5 mg, administered 3 to 7 nights per week for 24 weeks, in patients with chronic primary insomnia: a 6-month, randomized, double-blind, placebo-controlled, parallel-group, multicenter study. Sleep. 2008; 31(1):79–90.

19. Hills M, Armitage P. The two-period cross-over clinical trial. Br J Clin Pharmacol. 1979; 8(1):7–20.

20. Roden DM, Tyndale RF. Pharmacogenomics at the tipping point: challenges and opportunities. Clin Pharmacol Ther. 2011; 89(3):323–7.

21. Davit BM, et al. Highly variable drugs: observations from bioequivalence data submitted to the FDA for new generic drug applications. AAPS J. 2008; 10(1): 148–156.

22. Koo CM, et al. Cannabidiol for treating Lennox-Gastaut Syndrome and Dravet Syndrome in Korea. J Korean Med Sci. 2020; 35(50): e427.

23. Koubeissi M. Anticonvulsant effects of cannabidiol in dravet syndrome. Epilepsy Curr. 2017; 17(5): 281–282.

24. Duan N, et al. Single-patient (n-of-1) trials: a pragmatic clinical decision methodology for patient-centered comparative effectiveness research. Journal of Clinical Epidemiology. 2013; 66(8 Suppl):S21–8.

25. 25. https://www.whoop.com/us/en/thelocker/average-sleep-s

26. Institute of Medicine (US) Committee on Sleep Medicine and Research; Colten HR, Altevogt BM, editors. Sleep disorders and sleep deprivation: an unmet public health problem. Washington (DC): National Academies Press (US); 2006. 2, Sleep Physiology.

27. Li J, et al. Sleep in Normal Aging. Sleep Med Clin. 2018; 13(1):1–11.

28. Ohayon MM, et al. Meta-analysis of quantitative sleep parameters from childhood to old age in healthy individuals: developing normative sleep values across the human lifespan. Sleep. 2004; 27(7):1255–73.

29. Leary EB, et al. Association of rapid eye movement sleep with mortality in middle-aged and older adults. JAMA Neurol. 2020; 77(10):1241–1251.

30. Sehgal A, Mignot E. Genetics of sleep and sleep disorders. Cell; 2011; 146(2):194–207.

31. Åkerstedt T, Wright, KP Jr. Sleep Loss and Fatigue in Shift Work and Shift Work Disorder. Sleep Med Clin. 2009; 4(2):257–271.

32. Leweke FM, et al. Cannabidiol enhances anandamide signaling and alleviates psychotic symptoms of schizophrenia. Transl Psychiatry. 2012; 2(3):e94.

33. Murillo-Rodríguez E, et al. Anandamide modulates sleep and memory in rats. Brain Res 1998; 812:270–4.

34. Russo EB. Taming THC: potential cannabis synergy and phytocannabinoid-terpenoid entourage effects. Br J Pharmacol. 2011; 163(7):1344–64.

35. Kaufmann R, et al. Observed impact of long-term consumption of oral cannabidiol on liver function in healthy adults. Cannabis Cannabinoid Res. 2023; 8(1):148–154.

36. Ried K, et al. Medicinal cannabis improves sleep in adults with insomnia: a randomised double-blind placebo-controlled crossover study. J Sleep Res. 2023; 32(3):e13793.

37. Feinberg I, et al. Effects of high dosage delta-9-tetrahydrocannabinol on sleep patterns in man. Clin Pharmacol Ther. 1975;17(4):458–66.

38. Lancel M. Role of GABAA receptors in the regulation of sleep: initial sleep responses to peripherally administered modulators and agonists. Sleep. 1999; 22(1):33–42.

39. De Crescenzo F, et al. Comparative effects of pharmacological interventions for the acute and long-term management of insomnia disorder in adults: a systematic review and network meta-analysis. Lancet. 2022; 400:170–184.

